# Germline mutations in young-onset sporadic pituitary macroadenomas: a multigene panel analysis

**DOI:** 10.1101/2024.06.02.24308129

**Authors:** Leonor M. Gaspar, Catarina I. Gonçalves, Ema L. Nobre, Fernando Fonseca, Cláudia Amaral, João S. Duarte, Luísa Raimundo, Catarina Saraiva, Luísa Cortez, Olinda Marques, Manuel C. Lemos

## Abstract

**Objective:** Mutations in several genes have been associated with familial forms of pituitary adenomas. Sporadic pituitary adenomas (i.e. with no family history or coexistent endocrine tumours) are also occasionally found to result from germline mutations in these genes, especially in young patients with larger tumours. The aim of this study was to determine the frequency of germline mutations in patients with young-onset sporadic pituitary macroadenomas.

**Methods:** A cohort of 225 Portuguese patients with sporadic pituitary macroadenomas diagnosed before the age of 40 years was studied by whole exome sequencing (WES) followed by the analysis of a virtual panel of 29 genes that have been associated with predisposition to pituitary adenomas.

**Results:** Pathogenic and likely pathogenic variants were identified in 16 (7.1%) of patients. The affected genes were *AIP* (n=4), *PMS2* (n=4), *MEN1* (n=2), *VHL* (n=2), *CDH23* (n=1), *MSH2* (n=1), *SDHB* (n=1), and *TP53* (n=1). In patients diagnosed under the ages of 30 and 18 years, the frequency of mutations increased to 9.0% and 12.0%, respectively.

**Conclusion:** This is so far the largest multigene analysis of patients with young-onset sporadic pituitary macroadenomas. We confirmed the *AIP* as the most frequently involved gene, but also uncovered rarer genetic causes of pituitary adenomas, including the first independent confirmation of a role of the *CDH23* gene. The results may contribute to a better understanding of the genetic landscape of these tumours and help to decide which genes to include in the genetic screening of patients with young-onset pituitary macroadenomas.

## Introduction

Most pituitary adenomas occur sporadically and are often attributed to acquired somatic and epigenetic mutations (1). However, a subset of cases arises within a familial context, either as part of syndromic diseases or as familial isolated pituitary adenomas (FIPA), which are caused by pathogenic germline mutations (2). Tumours within familial settings tend to be more aggressive, manifesting at a younger age, with larger sizes, increased invasiveness, and resistance to standard treatments (3). An expanding list of genes, including *AIP*, *CABLES1*, *CDH23*, *CDKN1A*, *CDKN1B*, *CDKN2B*, *CDKN2C*, *DICER1*, *GNAS*, *GPR101*, *MAX*, *MEN1*, *MLH1*, *MSH2*, *MSH6*, *NF1*, *PMS2*, *PRKACA*, *PRKACB*, *PRKAR1A*, *RET*, *SDHA*, *SDHAF2*, *SDHB*, *SDHC*, *SDHD*, *TP53*, *USP8*, and *VHL*, has been identified with germline or mosaic mutations predisposing individuals to pituitary adenomas (4–6). Identifying these genetic alterations is not only crucial for accurate diagnosis and personalized treatment, but also provides valuable insights into the molecular pathways disrupted in these tumours (7).

While sporadic cases traditionally lack a clear hereditary component, several studies have shown that a variable proportion of these cases harbour germline mutations (8). The most extensively studied gene is *AIP*, which was first associated with FIPA (9), but later found to be mutated in many apparently sporadic cases, particularly among patients of younger ages and with larger tumours (10).

We recently screened a cohort of patients diagnosed with young-onset sporadic pituitary macroadenomas for *AIP* mutations (11). This revealed the presence of *AIP* mutations in 1.8%, 3.4% and 5.0% of patients diagnosed under the ages of 40, 30 and 18 years, respectively (11). Building upon this, we have now employed next-generation sequencing to expand the genetic screening to all genes with germline mutations that have so far been associated with familial isolated or syndromic pituitary adenomas.

## Materials and Methods

### Subjects

This was a follow-up study of a Portuguese multicentre cohort that had been previously studied by conventional (Sanger) sequencing of the *AIP* gene (11). A total of 225 patients were available for this study. Inclusion criteria were patients with macroadenomas (tumour greater diameter ≥ 1 cm) diagnosed under the age of 40 years. Exclusion criteria were patients with a family history of pituitary adenomas (i.e. affected first or second degree family member) or with evidence of a syndromic form of pituitary adenoma. Mean age (± standard deviation) at diagnosis was 29.1 ± 7.3 years, 122 patients were under 30 years at diagnosis, and 25 patients were under 18 years at diagnosis. Gender distribution was 116 (51.6%) females and 109 (48.4%) males. Tumour classification was based on histological examination or, in the case of prolactinomas, by clinical, hormonal and radiological examination. Eighty-one (36.0%) patients had prolactinomas, 62 (27.6%) had somatotrophinomas, 37 (16.4%) had non-functioning pituitary adenomas, 16 (7.1%) had mixed-secreting pituitary adenomas, 15 (6.7%) had corticotrophinomas, seven (3.1%) had gonadotrophinomas, one (0.4%) had a thyrotrophinoma, and six (2.7%) had adenomas with undetermined histology. The control population comprised 298 Portuguese individuals (50 healthy blood donors and 248 patients with unrelated disorders). The study was approved by the Ethics Committee of the Faculty of Health Sciences, University of Beira Interior (Ref: CE-UBI-Pj-2018-027 and CE-FCS-2011-003) and written informed consent was obtained from all subjects.

### Whole exome sequencing (WES) and virtual gene panel

Genomic deoxyribonucleic acid (DNA) was extracted from the peripheral blood leukocytes of each patient and used for WES analysis according to previously described methods (12). A virtual gene panel was created, consisting of 29 genes in which germline or mosaic mutations have been reported in patients with familial isolated or syndromic pituitary adenomas (4–6), namely *AIP* (NM_003977.3), *CABLES1* (NM_001100619.2), *CDH23* (NM_022124.5), *CDKN1A* (NM_078467.3), *CDKN1B* (NM_004064.4), *CDKN2B* (NM_004936.4), *CDKN2C* (NM_001262.3), *DICER1* (NM_177438.3)*, GNAS* (NM_000516.7)*, GPR101* (NM_054021.1)*, MAX* (NM_002382)*, MEN1* (NM_130799.2)*, MLH1* (NM_000249.4), *MSH2* (NM_000251.3)*, MSH6* (NM_000179.3)*, NF1* (NM_000267.3)*, PMS2* (NM_000535.7), *PRKACA* (NM_002730.4), *PRKACB* (NM_182948.4)*, PRKAR1A* (NM_002734.5)*, RET* (NM_020975.4)*, SDHA* (NM_004168.3)*, SDHAF2* (NM_017841.2)*, SDHB* (NM_003000.3)*, SDHC* (NM_003001.5)*, SDHD* (NM_003002.3)*, TP53* (NM_000546.6), *USP8* (NM_005154.5), and *VHL* (NM_000551.3).

### Interpretation of genetic variants

Genetic variants were filtered according to the following cumulative criteria: 1) Location in one of the 29 genes previously implicated in pituitary adenomas; 2) Location in coding transcripts used by the Human Genome Mutation Database (HGMD) (13); 3) Location in coding exons or up to ten nucleotides adjacent to the coding exons; and 4) Population allele frequency less than 0.001 in the Genome Aggregation Database (gnomAD) and 1000 Genomes database (14). The variants selected by these criteria were classified as benign (B), likely benign (LB), variant of uncertain significance (VUS), likely pathogenic (LP) or pathogenic (P), according to American College of Medical Genetics and Genomics (ACMG) criteria (15) and ClinGen recommendations (16), using a web-based variant interpretation tool (Franklin by Genoox, reference hg19, https://franklin.genoox.com/, accessed on 30 March 2024). Filtered variants were screened in an in-house database of 298 Portuguese control individuals to assess the possibility of variants being population-specific common polymorphisms.

### Validation of genetic variants by Sanger sequencing

Variants classified as pathogenic and likely pathogenic were confirmed by conventional Sanger sequencing using a semi-automated DNA sequencer (STAB VIDA, Caparica, Portugal; and ABI 3730XL, Applied Biosystems; Thermo Fisher Scientific, Waltham, MA, USA).

## Results

### Rare sequence variants identified in the 29 analysed genes

A total of 154 (141 different) rare sequence variants (population allele frequency <0.001) were identified in 114 of the 225 patients. These variants were found in 25 of the 29 analysed genes and included three pathogenic, 13 likely pathogenic (11 different), 63 VUS (56 different), 64 likely benign (61 different) and 11 benign (10 different) (Supplemental Data 1). All rare sequence variants were identified in the heterozygous state.

### Pathogenic and likely pathogenic variants

Pathogenic and likely pathogenic variants were identified in 16 (7.1%) patients with young-onset sporadic pituitary macroadenomas. These consisted of four *AIP* gene mutations (previously reported by us (11)) (p.Ser53ThrfsTer36, p.Arg81Ter, p.Leu115TrpfsTer41, and p.Glu246Ter), four *PMS2* mutations (three patients with p.Asn335Ser, and one with p.Asp486GlufsTer109), two *MEN1* mutations (p.Trp183Ter, and p.Arg314_Asp315del), two *VHL* mutations (p.Lys196Glu, and p.Glu52Ter), one *CDH23* mutation (p.Glu2520Lys), one *MSH2* mutation (p.Arg524His), one *SDHB* mutation (p.Ile127Leu), and one *TP53* mutation (p.Arg282Gln) (Table 1 and Figure 1).

**Figure 1.**
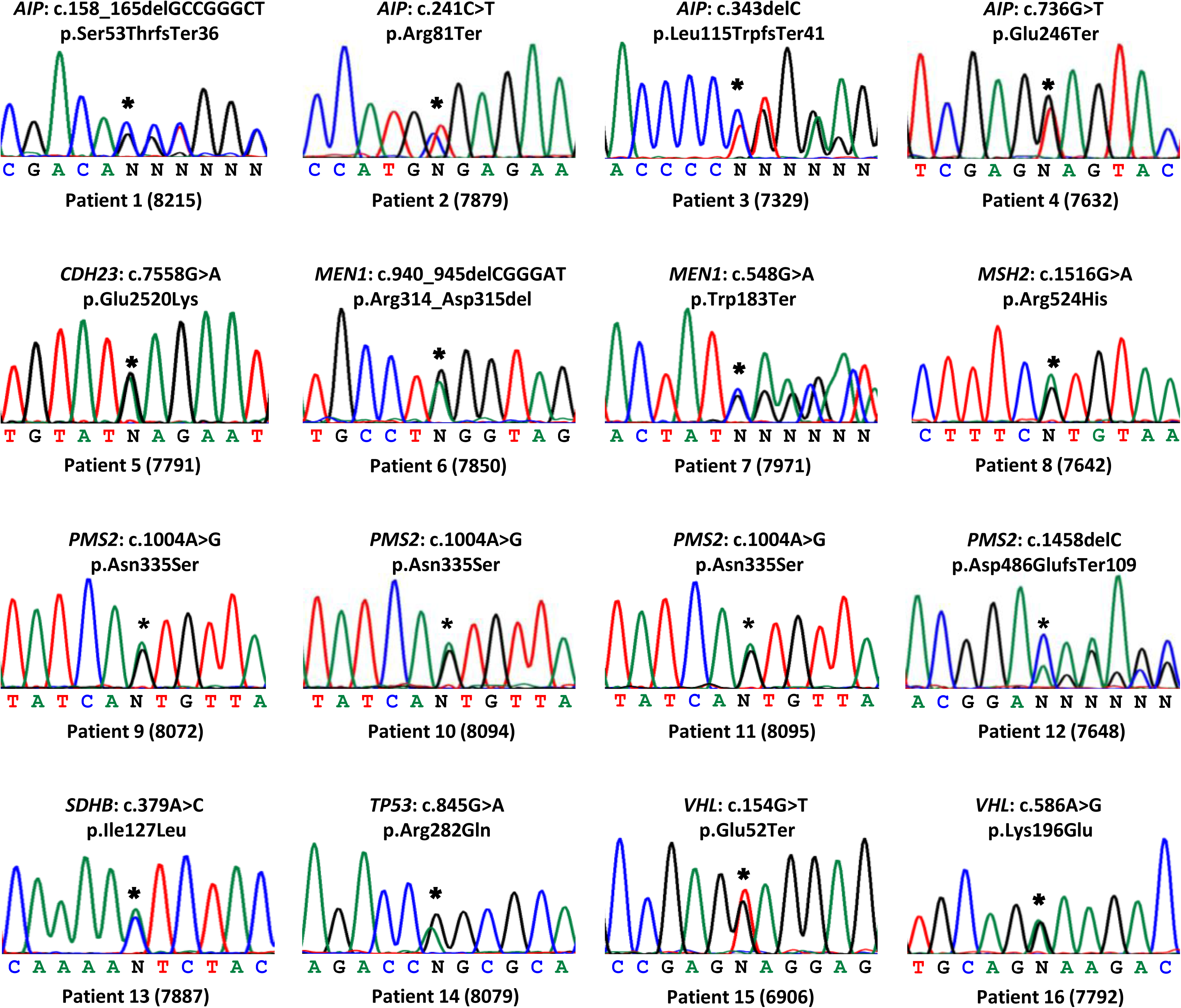
Germline mutations identified in patients. The Sanger sequencing chromatograms are presented for each heterozygous mutation (indicated by an asterisk) and surrounding nucleotides.

**Table 1.** Clinical and genetic characteristics of 16 patients with pathogenic (P) and likely pathogenic (LP) variants.

| Gene<br>(transcript) | Patient<br>number<br>(id) | Sex | Age at<br>diagnosis<br>(yr) | Type of<br>adenoma<br>(hormones<br>produced) | Size of<br>adenoma<br>(mm) | Variant (nucleotide change,<br>protein change) (a) | Effect | Allele<br>frequency in<br>GnomAD | Allele<br>frequency in<br>Portuguese<br>controls | ACMG<br>classification<br>(criteria) (b) | Previous<br>report |
| --- | --- | --- | --- | --- | --- | --- | --- | --- | --- | --- | --- |
| *AIP*<br>(NM_003977.3) | 1 (8215) | F | 19-30 | GH | 20 | c.158_165delGCCGGGCT,<br>p.Ser53ThrfsTer36 | Frameshift<br>deletion | 0 | 0 | LP (PVS1, PM2) | (11)* |
|  | 2 (7879) | M | 19-30 | GH | 26 | c.241C>T, p.Arg81Ter | Nonsense | 0 | 0 | P (PVS1, PM2,<br>PS4) | (49) (11)* |
|  | 3 (7329) | M | ≤18 | GH/PRL | 14 | c.343delC,<br>p.Leu115TrpfsTer41 | Frameshift<br>deletion | 0 | 0 | LP (PVS1, PM2) | (50) (11)* |
|  | 4 (7632) | F | 19-30 | GH | 28 | c.736G>T, p.Glu246Ter | Nonsense | 0 | 0 | LP (PVS1, PM2) | (11)* |
| *CDH23*<br>(NM_022124.5) | 5 (7791) | M | 19-30 | GH | 25 | c.7558G>A, p.Glu2520Lys | Missense | 0 | 0 | LP (PS4, PM2,<br>PP3, PP5) | (34) |
| *MEN1*<br>(NM_130799.2) | 6 (7850) | M | 31-40 | GH/PRL | 60 | c.548G>A, p.Trp183Ter | Nonsense | 0 | 0 | P (PVS1, PS4,<br>PP5, PM2) | (26) |
|  | 7 (7971) | F | 19-30 | GH/PRL/TSH | 40 | c.940_945delCGGGAT,<br>p.Arg314_Asp315del | In-frame<br>deletion | 0 | 0 | LP (PM1, PM2,<br>PM4) | None |
| *MSH2*<br>(NM_000251.3) | 8 (7642) | F | 19-30 | PRL | >10 | c.1571G>A, p.Arg524His | Missense | 0.000011 | 0 | LP (PM2, PM5,<br>PP3) | (37) |
| *PMS2*<br>(NM_000535.7) | 9 (8072) | M | 31-40 | GH | >10 | c.1004A>G, p.Asn335Ser | Missense | 0.000273 | 0 | LP (PP3, PM2,<br>BP6) | (24) |
|  | 10 (8094) | F | 31-40 | PRL | 31 | c.1004A>G, p.Asn335Ser | Missense | 0.000273 | 0 | LP (PP3, PM2,<br>BP6) | (24) |
|  | 11 (8095) | F | 31-40 | Non-functioning | 40 | c.1004A>G, p.Asn335Ser | Missense | 0.000273 | 0 | LP (PP3, PM2,<br>BP6) | (24) |
|  | 12 (7648) | M | 19-30 | PRL | >20 | c.1458delC,<br>p.Asp486GlufsTer109 | Frameshift<br>deletion | 0 | 0 | LP (PVS1, PM2) | None |
| *SDHB*<br>(NM_003000.3) | 13 (7887) | F | 19-30 | PRL | 20 | c.379A>C, p.Ile127Leu | Missense | 0.000004 | 0 | LP (PM1, PM2,<br>PM5, PP2, PP3) | (41) |
| *TP53*<br>(NM_000546.6) | 14 (8079) | F | 31-40 | ACTH | 14 | c.845G>A, p.Arg282Gln | Missense | 0.000004 | 0 | LP (BS3, PM2,<br>PM5, PM1, PP3,<br>PP5) | (44) |
| *VHL*<br>(NM_000551.3) | 15 (6906) | F | ≤18 | PRL | >10 | c.154G>T, p.Glu52Ter | Nonsense | 0.000018 | 0 | LP (PVS1, PM2) | (30) |
|  | 16 (7792) | M | ≤18 | TSH | 10 | c.586A>G, p.Lys196Glu | Missense | 0 | 0 | P (PM1, PM2,<br>PM3, PP2, PP3,<br>PP5) | (31) |

id, identification (anonymization code not known to anyone outside the research group); F, female; M, male; yr, years (presented as age range to protect patient privacy); FSH, follicle stimulating hormone; GH, growth hormone; LH, luteinizing hormone; PRL, prolactin; TSH, thyroid stimulating hormone; mm, millimeters; GnomAD, Genome Aggregation Database (v2.1.1). (a) All variants were heterozygous. (b) American College of Medical Genetics and Genomics (ACMG) classification of variants (P, pathogenic; LP, likely pathogenic) was based on the evidence for pathogenicity (very strong (PVS1), moderate (PM1–6), or supporting (PP1–5)). ACMG classifications were based on the web-based variant interpretation tool Franklin (Genoox Ltd, https://franklin.genoox.com/), accessed on 30 March 2024. \*Publication by the authors that included the same patient.

### Prevalence of mutations according to age of diagnosis

The prevalence of mutations was higher in patients with younger ages at diagnosis. The prevalence of mutations in patients diagnosed up until the age of 40, 30 and 18 years was 7.1% (16/225), 9.0% (11/122), and 12.0% (3/25), respectively.

### Clinical characteristics of patients with mutations

The clinical characteristics of the 16 patients with identified mutations are presented in Table 1. Patients had no personal history of additional tumours or other syndromic features at the time of inclusion in the study. However, two patients with *MEN1* mutations were found to have hyperparathyroidism during or after undertaking the genetic studies.

## Discussion

Our analysis of 225 patients with young-onset sporadic pituitary macroadenomas showed that 16 (7.1%) patients had germline mutations in genes that are associated with familial forms of pituitary adenomas. These mutations involved the *AIP* (1.8% of patients), *PMS2* (1.8%), *MEN1* (0.9%), *VHL* (0.9%), *CDH23* (0.4%), *MSH2* (0.4%), *SDHB* (0.4%), and *TP53* (0.4%) genes.

The *AIP* gene is associated with FIPA (9), but has also been extensively studied in patients with sporadic pituitary adenomas. The prevalence of *AIP* germline mutations in patients with sporadic pituitary macroadenomas under the age of 40 has been reported to vary from 0% to 18% (11), depending of the country of origin, clinical characteristics of the cohort, and criteria used for the classification of genetic variants. We found four (1.8%) patients with *AIP* mutations, which were frameshift (p.Ser53ThrfsTer36, and p.Leu115TrpfsTer41) and nonsense (p.Arg81Ter, and p.Glu246Ter) mutations expected to lead to a premature stop codon and to the formation of a shorter protein or to nonsense-mediated decay (17). These *AIP* mutations were all found in patients with GH-secreting adenomas, in agreement with the higher prevalence of *AIP* mutations in this tumour type (18). These results confirm the results of our previous Sanger sequencing of the *AIP* gene in this cohort of patients (11).

The *PMS2* gene is associated with Lynch syndrome (19), which is characterised by the occurrence of a variety of tumours that include colorectal, endometrial, ovarian and gastric cancers (20). Although some cases of aggressive pituitary tumours have been reported in patients with Lynch syndrome (21–23), the prevalence of *PMS2* mutations in sporadic pituitary adenomas has never been reported before. We found four (1.8%) patients with *PMS2* mutations, with no other apparent manifestations of Lynch syndrome. These consisted of a previously reported (24) missense (p.Asn335Ser) mutation that was identified in three unrelated patients, diagnosed with a somatotrophinoma, prolactinoma and non-functioning pituitary adenoma, and a novel frameshift (p.Asp486GlufsTer109) mutation in a patient with a prolactinoma. Thus, our study suggests that the *PMS2* gene has a more important role in pituitary tumorigenesis than previously acknowledged.

The *MEN1* gene is associated with the multiple endocrine neoplasia type 1 (MEN1) syndrome, which is characterised by the occurrence of parathyroid, pancreatic and pituitary tumours (25, 26). *MEN1* mutations are occasionally found in patients with pituitary adenomas without other MEN1 manifestations. A previous study identified *MEN1* mutations in 3.4% of patients with sporadic pituitary macroadenomas diagnosed before the age of 30 (27). Our study found two (0.9%) patients with *MEN1* mutations, which consisted of a previously reported (26) nonsense mutation (p.Trp183Ter) and a novel in-frame deletion (p.Arg314_Asp315del). Both patients had mixed GH-secreting adenomas. It is interesting to note that although there were no other apparent manifestations of the MEN1 syndrome at the time of the diagnosis of the pituitary adenoma, both patients were eventually found to have hyperparathyroidism during or after undertaking the genetic studies.

The *VHL* gene is associated with the Von Hippel–Lindau (VHL) syndrome, which is characterised by tumours in several organs, such as retinal and central nervous system haemangioblastomas, pheochromocytomas and clear-cell renal carcinomas (28). Pituitary adenomas have also been described in patients with the VHL syndrome (29). However, the prevalence of *VHL* mutations in sporadic pituitary adenomas has not been reported. We found two (0.9%) patients with *VHL* mutations, with no other apparent manifestations of the VHL syndrome. These consisted of a previously reported (30) nonsense (p.Glu52Ter) mutation in a patient with a prolactinoma and a previously reported (31) missense (p.Lys196Glu) mutation in a patient with a thyrotrophinoma. The latter mutation was previously reported in homozygosity in a patient with autosomal recessive congenital erythrocytosis, but with no evidence of the VHL syndrome (31). Therefore, it remains to be clarified if heterozygosity for this particular mutation, as found in our patient, can cause the VHL syndrome.

The *CDH23* gene is associated with the autosomal recessive Usher syndrome, which is characterized by congenital deafness (32). However, a study by Zhang et al. (33) demonstrated the presence of *CDH23* heterozygous mutations in 33% and 12% of familial and isolated pituitary adenomas, respectively. So far, these results have not been independently confirmed. Furthermore, there have been no reports of a higher incidence of pituitary adenomas in patients with Usher syndrome or in their heterozygous relatives. We found one (0.4%) patient with a GH-secreting adenoma and a previously reported (34) *CDH23* missense mutation (p.Glu2520Lys). Thus, our study represents the first independent confirmation of a role of *CDH23* in the development of pituitary adenomas.

The *MSH2* gene is also associated with Lynch syndrome (35, 36) and some affected patients have been reported to have aggressive pituitary adenomas (21–23). We found one (0.4%) patient with a previously reported (37) *MSH2* missense mutation (p.Arg524His), who had a prolactinoma with no other apparent manifestations of Lynch syndrome.

The *SDHB* gene is associated with paragangliomas and pheochromocytomas (38). Pituitary adenomas occasionally occur in association with these (3PA, Phaeochromocytoma, Paraganglioma and Pituitary adenoma association) (39). However, the prevalence of *SDHB* mutations in sporadic pituitary adenomas is unknown. A French study of 263 patients with sporadic pituitary adenomas revealed two mutations in the *SDHA* gene, one in the *SDHC* gene, but none in *SDHB* (40). We found one (0.4%) patient with a previously reported (41) *SDHB* missense mutation (p.Ile127Leu), who had a prolactinoma without any other syndromic manifestations.

The *TP53* gene is considered the most mutated tumour suppressor gene in human cancers (42). Germline mutations in this gene are associated with the Li-Fraumeni syndrome, which predisposes to soft tissue sarcomas, osteosarcoma, breast cancer, leukaemia, and adrenocortical carcinoma (43). The role of the *TP53* gene in pituitary adenomas is less clear. The *TP53* gene was included in our gene panel because a recent review (6) listed it as one of the genes in which germline mutations are implicated in pituitary adenomas. However, although somatic mutations in *TP53* have been reported in pituitary adenomas (1), we found no reports of germline mutations in patients with these tumours. Nevertheless, in our study, we found one (0.4%) patient with a previously reported (44) *TP53* germline missense mutation (p.Arg282Gln), who had a corticotrophinoma without any other syndromic manifestations. Thus, our study reports for the first time a *TP53* germline pathogenic mutation in a patient with a pituitary adenoma.

None of the patients included in our study had other coexistent tumours or syndromic features or family history that would raise the suspicion of a germline mutation. The unexpected identification of germline mutations in a subset of patients with sporadic tumours could have several explanations. First, family history was self-reported by the patients and may have been inaccurate or incomplete. Second, the lack of other affected family members could have been due to incomplete penetrance of the mutation or to a *de novo* mutation in the patient. Third, other syndromic manifestations could have been missed on clinical screening or absent due to variable expression of the mutation. Importantly, our identification of patients with germline mutations will improve their clinical management, allow the screening of additional syndromic manifestations, and allow the identification of additional affected family members that can be screened for the disorder (6).

Previous studies of sporadic pituitary adenomas have mainly focused on the *AIP* gene, as this is the most commonly mutated gene in such cases (8). Only three other studies performed gene panel analyses in patients with sporadic pituitary adenomas, but with a limited number of genes (≤ 9) that did not include for example the *VHL, PMS2* or *CDH23* genes (40, 45, 46). Nevertheless, these studies were able to identify pathogenic variants in 3.8% to 10% of patients with young-onset sporadic pituitary adenomas.

In our study, we analysed the largest panel of genes so far in patients with sporadic pituitary adenomas. We confirmed the *AIP* as the most frequently involved gene in these patients, but also uncovered rarer genetic causes of pituitary adenomas. Altogether, germline mutations were present in 7.1% of our patients diagnosed with sporadic macroadenomas under the age of 40 years. However, this proportion increased to 9.0% and 12.0%, in patients diagnosed under the ages of 30 and 18 years, respectively. This is in agreement with the general observation that tumours arising in younger ages are more likely to have a genetic cause.

The existence of subsets of patients at higher risk of harbouring germline mutations has led to recommendations for *AIP* and *MEN1* mutation testing in patients with pituitary macroadenomas diagnosed under the age of 30 (10, 27) or 40 years (47). However, there are currently no recommendations for additional genetic testing of sporadic pituitary adenomas that have been shown to be *AIP* and *MEN1* mutation-negative. Our study suggests that testing such patients for a wider gene panel may uncover further cases of genetically-determined pituitary adenomas.

Our study has some limitations. First, we did not look for copy number variants or mutations in non-coding genomic regions. Second, we did not look for mutations in other genes beyond those that have so far been associated with pituitary adenomas. Third, we found a large number of VUS, for which there is currently insufficient evidence for an association with the disorder, but that may need reclassification over time (48).

In conclusion, we found a prevalence of 7.1% germline mutations in patients with young-onset pituitary macroadenomas. These include mutations in the *AIP*, *MEN1*, *MSH2*, *PMS2*, *SDHB*, *TP53* and *VHL* genes and the first independent confirmation of a mutation in the *CDH23* gene. Our results may contribute to a better understanding of the genetic landscape of these tumours and help to decide which genes to include in the genetic screening of patients with young-onset pituitary macroadenomas.

## Supplementary data

Supplementary data 1. Rare sequence variants identified in patients with young-onset sporadic pituitary macroadenomas.

## Conflict of interest

The authors declare that there is no conflict of interest that could be perceived as prejudicing the impartiality of the research reported.

## Funding

This research was funded by the Portuguese Foundation for Science and Technology (FCT, project grants PTDC/MEC-MET/29489/2017 and UIDB/00709/2020). LM Gaspar was the recipient of a PhD fellowship from FCT (SFRH/BD/147160/2019).

## Acknowledgements

The authors are grateful to the following clinicians for collecting patient samples and clinical data: Ana Agapito (Lisboa), Ana Monteiro (Braga), Ana Palha (Lisboa), Bernardo Pereira (Ponta Delgada), Daniela Cavaco (Lisboa), Davide Carvalho (Porto), Hélder Simões (Lisboa), Henrique Luiz (Almada), Inês Barros (Braga), Inês Damásio (Lisboa), Isabel Inácio (Porto), Joana Pereira (Lisboa), João Anselmo (Ponta Delgada), Maria João Bugalho (Lisboa), Maria Manuel Silva (Porto), Maria Salomé (Lisboa), Mariana Barbosa (Braga), Rita Santos (Lisboa), Sara Donato (Lisboa), Sara Pinheiro (Lisboa), Sílvia Paredes (Braga), Teresa Martins (Coimbra), Teresa Rego (Lisboa), Tiago Silva (Lisboa), and Valeriano Leite (Lisboa).

## Data availability statement

The data from this study are available from the corresponding author upon reasonable request.

## Author contribution statement

LM Gaspar and CI Gonçalves contributed to the acquisition, analysis and curation of genetic data, and writing of the original draft of the manuscript. E Nobre, F Fonseca, C Amaral, J Sequeira Duarte, L Raimundo, C Saraiva, L Cortez, and O Marques contributed to the recruitment and clinical studies of the patients. MC Lemos contributed to the conceptualization, funding acquisition, resources and supervision of the project. All authors contributed to reviewing and editing and approved the final version of the manuscript.

## Notes

### Competing Interest Statement

The authors have declared no competing interest.

### Author Declarations

Ethics Committee of the Faculty of Health Sciences, University of Beira Interior gave ethical approval for this work (Ref: CE-UBI-Pj-2018-027 and CE-FCS-2011-003)

