## Supplemental Data 1 for "Germline mutations in young-onset sporadic pituitary macroadenomas: a multigene panel analysis"

### **Table S1.** Rare sequence variants identified in patients with young-onset sporadic pituitary macroadenomas

| Gene (transcript) | Patient number (id) | Sex | Age at diagnosis (yr) | Type of adenoma (hormones produced) | Size of adenoma (mm) | Variant (nucleotide change, protein change) (a) | Effect | Allele frequency in GnomAD | Allele frequency in Portuguese controls | ACMG classification (criteria) (b) | Previous report (PMID) |
|---|---|---|---|---|---|---|---|---|---|---|---|
| *AIP* (NM_003977.3) | 8215 | F | 19-30 | GH | 20 | c.158_165delGCCGGGCT, p.Ser53ThrfsTer36 | Frameshift deletion | 0 | 0 | LP (PVS1, PM2) | 37149543* |
| | 7879 | M | 19-30 | GH | 26 | c.241C>T, p.Arg81Ter | Nonsense | 0 | 0 | P (PVS1, PM2, PS4) | 18381572, 37149543* |
| | 7329 | M | ≤18 | GH/PRL | 14 | c.343delC, p.Leu115TrpfsTer41 | Frameshift deletion | 0 | 0 | LP (PVS1, PM2) | 30851160, 37149543* |
| | 8458 | M | 19-30 | GH/PRL | 22 | c.695C>T, Pro232Leu | Missense | 0.000025 | 0 | VUS (PM2) | - |
| | 7632 | F | 19-30 | GH | 25 | c.736G>T, p.Glu246Ter | Nonsense | 0 | 0 | LP (PVS1, PM2) | 37149543* |
| | 8092 | F | 19-30 | GH | >10 | c.753G>A, p.Leu251Leu | Synonymous | 0.000171 | 0.0034 | LB (BP6, BP7) | 26792934 |
| | 8343 | F | 19-30 | GH | >10 | c.753G>A, p.Leu251Leu | Synonymous | 0.000171 | 0.0034 | LB (BP6, BP7) | 26792934 |
| | 7995 | F | 19-30 | PRL | 12 | c.896C>T, p.Ala299Val | Missense | 0.000544 | 0 | VUS (BP6) | 17360484 |
| | 8079 | F | 31-40 | ACTH | 14 | c.896C>T, p.Ala299Val | Missense | 0.000544 | 0 | VUS (BP6) | 17360484 |
| | 8158 | F | 31-40 | ACTH | 14 | c.896C>T, p.Ala299Val | Missense | 0.000544 | 0 | VUS (BP6) | 17360484 |
| | 8182 | F | 19-30 | PRL | 15 | c.896C>T, p.Ala299Val | Missense | 0.000544 | 0 | VUS (BP6) | 17360484 |
| *CABLES1* (NM_001100619.2) | 8462 | M | 19-30 | Non-functioning | >10 | c.17C>G, p.Ala6Gly | Missense | 0 | 0 | VUS (PM2, BP4) | - |
| | 7637 | F | 19-30 | GH | 34 | c.19G>C, p.Ala7Pro | Missense | 0 | 0 | VUS (PM2, BP4) | - |
| | 8163 | M | 19-30 | GH | 29 | c.19G>C, p.Ala7Pro | Missense | 0 | 0 | VUS (PM2, BP4) | - |
| | 7813 | F | 31-40 | GH | 40 | c.440C>T, p.Pro147Leu | Missense | 0.000159 | 0.0017 | VUS (PM2, BP4) | - |

| Gene (transcript) | Patient number (id) | Sex | Age at diagnosis (yr) | Type of adenoma (hormones produced) | Size of adenoma (mm) | Variant (nucleotide change, protein change) (a) | Effect | Allele frequency in GnomAD | Allele frequency in Portuguese controls | ACMG classification (criteria) (b) | Previous report (PMID) |
|---|---|---|---|---|---|---|---|---|---|---|---|
| | 7703 | M | 31-40 | Non-functioning | 30 | c.1050A>G, p.Ile350Met | Missense | 0.000004 | 0 | VUS (PM2, BP4) | - |
| | 8215 | F | 19-30 | GH | 20 | c.1260C>T, p.Ser420Ser | Synonymous | 0.000064 | 0.0034 | VUS (PM2) | - |
| | 8054 | F | 31-40 | PRL | 29 | c.1262C>G, p.Ser421Cys | Missense | 0.000004 | 0 | VUS (PM2) | - |
| *CDH23* (NM_022124.5) | 7820 | F | 31-40 | PRL | 20 | c.415G>A, p.Val139Ile | Missense | 0.000082 | 0.0034 | VUS (PM2) | 24416283 |
| | 7804 | M | 31-40 | GH | 17 | c.535G>A, p.Ala179Thr | Missense | 0.000020 | 0 | LB (PM2, BP4, BP6) | 30033219 |
| | 6905 | M | ≤18 | GH/PRL | >10 | c.1122G>T, p.Val374Val | Synonymous | 0.000028 | 0 | LB (PM2, BP6, BP7) | - |
| | 7686 | M | 19-30 | Non-functioning | >10 | c.1122G>T, p.Val374Val | Synonymous | 0.000028 | 0 | LB (PM2, BP6, BP7) | - |
| | 7826 | M | ≤18 | PRL | >10 | c.1450-10G>A | Intronic | 0.000053 | 0 | LB (PM2, BP4, BP6) | - |
| | 7631 | M | 19-30 | GH | 30 | c.1672G>A, p.Val558Met | Missense | 0.000107 | 0 | VUS (PM2, BP4) | - |
| | 7833 | M | 19-30 | PRL | 24 | c.1963G>A, p.Val655Ile | Missense | 0.000085 | 0.0017 | VUS (PM2) | - |
| | 7959 | M | 31-40 | FSH/LH | 23 | c.2836G>A, p.Val946Met | Missense | 0.000016 | 0 | VUS (PM2) | - |
| | 8215 | F | 19-30 | GH | 20 | c.2836G>A, p.Val946Met | Missense | 0.000016 | 0 | VUS (PM2) | - |
| | 7827 | M | 19-30 | GH/PRL | 11 | c.3231T>G, p.Pro1077Pro | Synonymous | 0.000122 | 0 | LB (PM2, BP6, BP7) | 12075507 |
| | 7853 | F | 19-30 | PRL | 10 | c.3739C>T, p.Arg1247Cys | Missense | 0.000207 | 0 | VUS (PM2) | - |
| | 8259 | F | 19-30 | GH | >10 | c.3852G>A, p.Ser1284Ser | Synonymous | 0.000143 | 0 | LB (BS1, BP6, BP7) | - |
| | 7878 | M | 31-40 | Undefined | 16 | c.3986G>A, p.Gly1329Asp | Missense | 0.000300 | 0 | VUS (PP3) | - |
| | 8130 | F | 31-40 | ACTH | 14 | c.4359+9C>A | Intronic | 0.000032 | 0 | VUS (PM2, BP6) | - |

| Gene (transcript) | Patient number (id) | Sex | Age at diagnosis (yr) | Type of adenoma (hormones produced) | Size of adenoma (mm) | Variant (nucleotide change, protein change) (a) | Effect | Allele frequency in GnomAD | Allele frequency in Portuguese controls | ACMG classification (criteria) (b) | Previous report (PMID) |
|---|---|---|---|---|---|---|---|---|---|---|---|
| | 7986 | M | 19-30 | PRL | 40 | c.4582G>A, p.Glu1528Lys | Missense | 0.000132 | 0 | VUS (PM2, BP4) | - |
| | 8130 | F | 31-40 | ACTH | 14 | c.4621G>A, p.Val1541Met | Missense | 0.000101 | 0 | VUS | 25257991 |
| | 8204 | M | 31-40 | GH | >10 | c.4780C>T, p.Arg1594Cys | Missense | 0.000121 | 0 | VUS (PM2, PP3) | - |
| | 7889 | M | 31-40 | PRL | >40 | c.5050C>T, p.Arg1684Cys | Missense | 0.000453 | 0 | VUS (BP4) | - |
| | 7745 | F | 31-40 | Non-functioning | 22 | c.5277G>A, p.Pro1759Pro | Synonymous | 0.000020 | 0 | LB (PM2, BP6, BP7) | - |
| | 7868 | F | 31-40 | GH | 18 | c.5626A>G, p.Ser1876Gly | Missense | 0 | 0 | VUS (PM2, BP4) | - |
| | 7986 | M | 19-30 | PRL | 40 | c.5685C>T, p.Arg1895Arg | Synonymous | 0.000008 | 0 | LB (PM2, BP6, BP7) | 19375528 |
| | 8204 | M | 31-40 | GH | >10 | c.6168C>T, p.Leu2056Leu | Synonymous | 0.000103 | 0 | LB (PM2, BP6, BP7) | - |
| | 7893 | F | 31-40 | Non-functioning | 22 | c.6264C>A, p.Val2088Val | Synonymous | 0.000020 | 0 | LB (PM2, BP6, BP7) | - |
| | 8152 | F | 19-30 | GH | >10 | c.6678C>T, p.Asn2226Asn | Synonymous | 0.000106 | 0 | LB (PM2, BP6, BP7) | - |
| | 7837 | M | 31-40 | PRL | 33 | c.6713-8G>A | Intronic | 0.000517 | 0 | LB (BS1, BP4, BP6) | 28912962 |
| | 7791 | M | 19-30 | GH | 25 | c.7558G>A, p.Glu2520Lys | Missense | 0 | 0 | LP (PS4, PM2, PP3, PP5) | 32467589 |
| | 7792 | M | ≤18 | TSH | 10 | c.8167G>C, p.Val2723Leu | Missense | 0.000346 | 0 | LB (BS1, BP4, BP6) | 22607986 |
| | 7632 | F | 19-30 | GH | 25 | c.8328C>T, p.Asn2776Asn | Synonymous | 0 | 0 | VUS (PM2) | - |
| | 7884 | F | 19-30 | GH | 18 | c.8407G>A, p.Val2803Ile | Missense | 0.000129 | 0 | VUS (BP4) | - |
| | 7904 | F | 19-30 | PRL | >10 | c.8577C>T, p.Ala2859Ala | Synonymous | 0.000019 | 0 | LB (PM2, BP6, BP7) | - |

| Gene (transcript) | Patient number (id) | Sex | Age at diagnosis (yr) | Type of adenoma (hormones produced) | Size of adenoma (mm) | Variant (nucleotide change, protein change) (a) | Effect | Allele frequency in GnomAD | Allele frequency in Portuguese controls | ACMG classification (criteria) (b) | Previous report (PMID) |
|---|---|---|---|---|---|---|---|---|---|---|---|
| | 8023 | M | 31-40 | FSH/LH | 31 | c.8914G>A, p.Glu2972Lys | Missense | 0.000050 | 0 | VUS (PM2, BP4) | - |
| | 8471 | M | 19-30 | PRL | 47 | c.9012T>G, p.Phe3004Leu | Missense | 0 | 0 | VUS (PM2) | - |
| | 8014 | F | 19-30 | PRL | 14 | c.9076C>T, p.Arg3026Trp | Missense | 0.000007 | 0 | VUS (PM2) | - |
| | 7688 | F | 19-30 | PRL | 12 | c.9078G>C, p.Arg3026Arg | Synonymous | 0.000004 | 0 | VUS (PM2) | 29048421 |
| | 7791 | M | 19-30 | GH | 25 | c.9750T>C, p.Thr3250Thr | Synonymous | 0.000025 | 0 | LB (PM2, BP6, BP7) | - |
| | 7634 | F | 19-30 | GH | >10 | c.9775C>T, p.His3259Tyr | Missense | 0.000012 | 0 | VUS (PM2, BP4) | - |
| | 8331 | M | 31-40 | PRL | 69 | c.9983G>A, p.Arg3328His | Missense | 0.000105 | 0 | LB (PM2, BS2, PP3, BP6) | - |
| | 7837 | M | 31-40 | PRL | 33 | c.10026C>T, p.Asp3342Asp | Synonymous | 0.000549 | 0 | LB (BS1, BP6, BP7) | 28912962 |
| *CDKN1B* (NM_004064.4) | 7636 | M | 19-30 | Non-functioning | 33 | c.471C>A, p.Thr157Thr | Synonymous | 0.000026 | 0 | LB (PM2, BP6, BP7) | - |
| | 8044 | F | 31-40 | PRL | 11 | c.492C>T, p.Asn164Asn | Synonymous | 0.000028 | 0.0017 | LB (PM2, BP6, BP7) | - |
| | 7689 | F | ≤18 | GH | >10 | c.589C>A, p.Gln197Lys | Missense | 0.000012 | 0 | VUS (PM2, BP1) | - |
| *CDKN2B* (NM_004936.4) | 7689 | F | ≤18 | GH | >10 | c.115G>A, p.Asp39Asn | Missense | 0.000049 | 0 | VUS (PM2, BP4) | - |
| *DICER1* (NM_177438.3) | 7911 | M | 31-40 | Undefined | >10 | c.21A>G, p.Gln7Gln | Synonymous | 0.000008 | 0 | LB (PM2, BP6, BP7) | - |
| | 8243 | F | 31-40 | ACTH | 10 | c.179C>T, p.Thr60Ile | Missense | 0.000050 | 0 | VUS (PM2, PP2, BP6) | 29399970 |
| | 7889 | M | 31-40 | PRL | >40 | c.1124C>G, p.Pro375Arg | Missense | 0.000329 | 0.0017 | VUS (PM2, PP2, BP6) | 29474644 |

| Gene (transcript) | Patient number (id) | Sex | Age at diagnosis (yr) | Type of adenoma (hormones produced) | Size of adenoma (mm) | Variant (nucleotide change, protein change) (a) | Effect | Allele frequency in GnomAD | Allele frequency in Portuguese controls | ACMG classification (criteria) (b) | Previous report (PMID) |
|---|---|---|---|---|---|---|---|---|---|---|---|
| | 8177 | M | 31-40 | PRL | 14 | c.1124C>G, p.Pro375Arg | Missense | 0.000329 | 0.0017 | VUS (PM2, PP2, BP6) | 29474644 |
| | 7680 | F | ≤18 | PRL | 12 | c.1293G>A, p.Glu431Glu | Synonymous | 0.000004 | 0 | LB (PM2, BP6, BP7) | - |
| | 7992 | M | 19-30 | PRL | 28 | c.2337A>G, p.Thr779Thr | Synonymous | 0.000046 | 0 | LB (PM2, BP6, BP7) | 29399970 |
| | 7971 | F | 19-30 | GH/PRL/FSH | 40 | c.5184C>A, p.Ser1728Ser | Synonymous | 0.000004 | 0 | LB (PM2, BP6, BP7) | - |
| | 7678 | F | 19-30 | PRL | 12 | c.5504A>G, p.Tyr1835Cys | Missense | 0.000080 | 0.0017 | VUS (PM2, PP2, PP3, BP6) | - |
| | 8342 | F | 31-40 | ACTH | 15 | c.5550C>G, p.Pro1850Pro | Synonymous | 0.000004 | 0 | LB (PM2, PP2, BP4, BP6) | - |
| *GNAS* (NM_000516.7) | 7675 | M | 19-30 | Non-functioning | 40 | c.18C>T, p.Asn6Asn | Synonymous | 0.000162 | 0 | LB (PM2, BP6, BP7) | - |
| | 7685 | F | 31-40 | GH | >10 | c.136C>T, p.Leu46Leu | Synonymous | 0.000048 | 0 | LB (PM2, BP6, BP7) | - |
| *GPR101* (NM_054021.1) | 8195 | F | 31-40 | GH | >10 | c.892G>A, p.Val298Ile | Missense | 0.000483 | 0 | LB (PM2, BS2, BP4, BP6) | - |
| | 8195 | F | 31-40 | GH | >10 | c.1170C>G, p.Pro390Pro | Synonymous | 0.000006 | 0 | VUS (PM2, BP7) | - |
| *MAX* (NM_002382.5) | 8115 | F | 19-30 | Non-functioning | 13 | c.*7C>T | 3' UTR | 0.000375 | 0 | LB (BP6, BP7) | - |
| *MEN1* (NM_130799.2) | 7850 | M | 31-40 | GH/PRL | 60 | c.548G>A, p.Trp183Ter | Nonsense | 0 | 0 | P (PVS1, PS4, PP5, PM2) | 9215690 |
| | 7971 | F | 19-30 | GH/PRL/TSH | 40 | c.940_945delCGGGAT, p.Arg314_Asp315del | In-frame deletion | 0 | 0 | LP (PM1, PM2, PM4) | - |

| Gene (transcript) | Patient number (id) | Sex | Age at diagnosis (yr) | Type of adenoma (hormones produced) | Size of adenoma (mm) | Variant (nucleotide change, protein change) (a) | Effect | Allele frequency in GnomAD | Allele frequency in Portuguese controls | ACMG classification (criteria) (b) | Previous report (PMID) |
|---|---|---|---|---|---|---|---|---|---|---|---|
| *MLH1* (NM_000249.4) | 8330 | F | 19-30 | ACTH | 11 | c.1080C>T, p.Ile360Ile | Synonymous | 0.000110 | 0 | LB (PM2, BP6, BP7) | - |
| | 8126 | F | 31-40 | GH | 22 | c.1608G>A, p.Gln536Gln | Synonymous | 0.000008 | 0 | LB (PM2, BP6, BP7) | - |
| | 7968 | M | 31-40 | PRL | 18 | c.198C>T, p.Thr66Thr | Synonymous | 0.000290 | 0 | LB (PM2, BP6, BP7) | 9833759 |
| | 7920 | M | ≤18 | ACTH | 41 | c.977T>C, p.Val326Ala | Missense | 0.000463 | 0 | VUS (PM2, BS2, PM5, PP3, BP6) | 8592341 |
| | 8095 | F | 31-40 | Non-functioning | 40 | c.1217G>A, p.Ser406Asn | Missense | 0.000903 | 0.0017 | VUS (PM2, BP6) | 9087566 |
| *MSH2* (NM_000251.3) | 7873 | F | 19-30 | PRL | 13 | c.1808C>G, p.Pro603Arg | Missense | 0.000156 | 0 | VUS (PM2, PP3, BP6) | 11726306 |
| | 7764 | F | 31-40 | PRL | 30 | c.725A>C, p.Asn242Thr | Missense | 0 | 0 | VUS (PM2, PP3) | - |
| | 8218 | M | 31-40 | Non-functioning | 25 | c.843A>T, p.Ser281Ser | Synonymous | 0.000032 | 0 | LB (PM2, BP6, BP7) | - |
| *MSH6* (NM_000179.3) | 7949 | M | 31-40 | GH | 25 | c.972G>A, p.Gln324Gln | Synonymous | 0.000076 | 0.0017 | LB (BP6, BP7) | 10777691 |
| | 7642 | F | 19-30 | PRL | >10 | c.1571G>A, p.Arg524His | Missense | 0.000011 | 0 | LP (PM2, PM5, PP3) | 29659569 |
| | 7728 | F | 31-40 | PRL | 12 | c.818G>T, p.Gly273Val | Missense | 0.000004 | 0.0017 | VUS (PM2, BP6) | 33827469 |
| | 7814 | F | ≤18 | PRL | 12 | c.840T>C, p.Ser280Ser | Synonymous | 0.000004 | 0 | LB (PM2, BP6, BP7) | - |
| | 7934 | F | 31-40 | PRL | >10 | c.3015A>G, p.Arg1005Arg | Synonymous | 0.000032 | 0 | LB (PM2, BP6, BP7) | - |

| Gene (transcript) | Patient number (id) | Sex | Age at diagnosis (yr) | Type of adenoma (hormones produced) | Size of adenoma (mm) | Variant (nucleotide change, protein change) (a) | Effect | Allele frequency in GnomAD | Allele frequency in Portuguese controls | ACMG classification (criteria) (b) | Previous report (PMID) |
|---|---|---|---|---|---|---|---|---|---|---|---|
| | 8128 | M | 31-40 | ACTH | 10 | c.3024C>T, p.Thr1008Thr | Synonymous | 0.000057 | 0 | LB (PM2, BP6, BP7) | - |
| | 7805 | F | 19-30 | Non-functioning | 35 | c.3260C>A, p.Pro1087His | Missense | 0.000099 | 0 | VUS (PP3, BP6) | 27398995 |
| | 7684 | F | 19-30 | PRL | 12 | c.3283C>T, p.Arg1095Cys | Missense | 0.000081 | 0 | VUS (PM2, PP3, BP6) | 28531214 |
| | 7845 | F | 31-40 | GH | >10 | c.3852G>A, p.Thr1284Thr | Synonymous | 0.000053 | 0.0034 | LB (PM2, BP6, BP7) | 14574004 |
| | 8340 | F | 31-40 | ACTH | >10 | c.3961A>G, p.Arg1321Gly | Missense | 0.000139 | 0 | VUS (PM2, PP3, BP6) | 16940983 |
| | 7703 | M | 31-40 | Non-functioning | 30 | c.3994T>C, p.Leu1332Leu | Synonymous | 0 | 0 | VUS (PM2, BP7) | - |
| | 8278 | M | 31-40 | FSH/LH | 41 | c.4002-10T>A | Intronic | 0.000257 | 0 | LB (PM2, BP4, BP6) | 21056691 |
| | 8342 | F | 31-40 | ACTH | 15 | c.4062G>T, p.Leu1354Leu | Synonymous | 0.000020 | 0 | LB (PM2, BP6, BP7) | - |
| *NF1* (NM_000267.3) | 7827 | M | 19-30 | GH/PRL | 11 | c.1176A>G, p.Gln392Gln | Synonymous | 0.000021 | 0 | LB (PM2, BP6, BP7) | - |
| | 7637 | F | 19-30 | GH | 34 | c.1186-4A>G | Intronic | 0.000033 | 0 | LB (PM2, BP4, BP6) | - |
| | 8093 | F | 31-40 | GH | 22 | c.1599C>G, p.Val533Val | Synonymous | 0.000156 | 0.0017 | LB (BS2, BP6, BP7) | - |
| | 8195 | F | 31-40 | GH | >10 | c.1599C>G, p.Val533Val | Synonymous | 0.000156 | 0.0017 | LB (BS2, BP6, BP7) | - |
| | 7770 | M | 31-40 | GH | >10 | c.3468C>T, p.Asn1156Asn | Synonymous | 0.000700 | 0.0050 | B (BS1, BP6) | 23460398 |
| | 8199 | F | 31-40 | PRL | >10 | c.3468C>T, p.Asn1156Asn | Synonymous | 0.000700 | 0.0050 | B (BS1, BP6) | 23460398 |

| Gene (transcript) | Patient number (id) | Sex | Age at diagnosis (yr) | Type of adenoma (hormones produced) | Size of adenoma (mm) | Variant (nucleotide change, protein change) (a) | Effect | Allele frequency in GnomAD | Allele frequency in Portuguese controls | ACMG classification (criteria) (b) | Previous report (PMID) |
|---|---|---|---|---|---|---|---|---|---|---|---|
| | 7632 | F | 19-30 | GH | 25 | c.4686A>G, p.Glu1562Glu | Synonymous | 0.000227 | 0 | LB (PM2, BP6, BP7) | 21354044 |
| | 8115 | F | 19-30 | Non-functioning | 13 | c.5694G>A, p.Glu1898Glu | Synonymous | 0.000127 | 0 | LB (PM2, BP6, BP7) | 33673681 |
| | 8319 | F | 19-30 | GH | >10 | c.7347T>C, p.Asn2449Asn | Synonymous | 0.000205 | 0 | B (BS1, BP6, BP7) | - |
| | 7958 | M | 31-40 | GH | >40 | c.7978A>G, p.Ile2660Val | Missense | 0.000173 | 0 | LB (PM2, PP2, BP4, BP6) | 16944272 |
| | 7675 | M | 19-30 | Non-functioning | 40 | c.8044C>T, p.Leu2682Leu | Synonymous | 0 | 0 | LB (PM2, BP6, BP7) | - |
| *PMS2* (NM_000535.7) | 8131 | F | 19-30 | Non-functioning | 22 | c.255G>A, p.Leu85Leu | Synonymous | 0.000182 | 0 | LB (PM2, BP6, BP7) | - |
| | 7850 | M | 31-40 | GH/PRL | 60 | c.383C>T, p.Ser128Leu | Missense | 0.000673 | 0 | B (BA1, BP6) | 20205264 |
| | 7850 | M | 31-40 | GH/PRL | 60 | c.830C>A, p.Thr277Lys | Missense | 0.000417 | 0 | B (BS1, BP6) | 17417778 |
| | 7835 | F | 19-30 | GH | 15 | c.831G>A, p.Thr277Thr | Synonymous | 0.000088 | 0 | LB (PM2, BP6, BP7) | - |
| | 8072 | M | 31-40 | GH | >10 | c.1004A>G, p.Asn335Ser | Missense | 0.000273 | 0 | LP (PP3, PM2, BP6) | 24549055 |
| | 8094 | F | 31-40 | PRL | 31 | c.1004A>G, p.Asn335Ser | Missense | 0.000273 | 0 | LP (PP3, PM2, BP6) | 24549055 |
| | 8095 | F | 31-40 | Non-functioning | 40 | c.1004A>G, p.Asn335Ser | Missense | 0.000273 | 0 | LP (PP3, PM2, BP6) | 24549055 |
| | 7648 | M | 19-30 | PRL | >20 | c.1458delC, p.Asp486GlufsTer109 | Frameshift deletion | 0 | 0 | LP (PVS1, PM2) | - |
| | 7833 | M | 19-30 | PRL | 24 | c.1533G>A, p.Thr511Thr | Synonymous | 0.000088 | 0 | LB (PM2, BP6, BP7) | 31159747 |

| Gene (transcript) | Patient number (id) | Sex | Age at diagnosis (yr) | Type of adenoma (hormones produced) | Size of adenoma (mm) | Variant (nucleotide change, protein change) (a) | Effect | Allele frequency in GnomAD | Allele frequency in Portuguese controls | ACMG classification (criteria) (b) | Previous report (PMID) |
|---|---|---|---|---|---|---|---|---|---|---|---|
| | 7633 | F | 19-30 | GH | >10 | c.1560G>C, p.Ala520Ala | Synonymous | 0 | 0 | LB (PM2, BP6, BP7) | - |
| *PRKAR1A* (NM_002734.5) | 8198 | M | 31-40 | PRL | 39 | c.101C>G, p.Ser34Cys | Missense | 0.000007 | 0 | VUS (PM2, PP2) | - |
| | 7868 | F | 31-40 | GH | 18 | c.156A>G, p.Glu52Glu | Synonymous | 0.000207 | 0 | LB (PM2, BP6, BP7) | - |
| | 7827 | M | 19-30 | GH/PRL | 11 | c.221G>A, p.Arg74His | Missense | 0.000393 | 0.0084 | B (PM5, PP2, BS1, BP6) | 29264456 |
| | 7839 | M | 19-30 | Non-functioning | 17 | c.662T>A, p.Val221Glu | Missense | 0 | 0 | VUS (PM2, PP2, PP3) | - |
| *RET* (NM_020975.4) | 8014 | F | 19-30 | PRL | 14 | c.96G>A, p.Ser32Ser | Synonymous | 0.000033 | 0 | LB (PM2, BP6, BP7) | 22174939 |
| | 8078 | M | 31-40 | PRL | 20 | c.785T>C, p.Val262Ala | Missense | 0.000248 | 0 | VUS (PM2, PP3, BP6) | 11955539 |
| | 8253 | M | ≤18 | PRL | 26 | c.1063+9G>A | Intronic | 0.000134 | 0 | VUS (PM2, BP6) | 7704557 |
| | 8051 | M | ≤18 | PRL | 47 | c.1462A>T, p.Thr488Ser | Missense | 0.000021 | 0.0017 | VUS (PM2, BP6) | 25425582 |
| | 8298 | M | 31-40 | Non-functioning | 20 | c.1529C>T, p.Ala510Val | Missense | 0.000248 | 0.0017 | B (BS1, BP6) | 20103606 |
| | 7850 | M | 31-40 | GH/PRL | 60 | c.1942G>A, p.Val648Ile | Missense | 0.000100 | 0.0050 | VUS (PM2, BP6) | 12466368 |
| | 7643 | M | 19-30 | Non-functioning | >10 | c.2052G>A, p.Pro684Pro | Synonymous | 0.000241 | 0.0017 | LB (BP6, BP7) | 28946813 |
| | 8057 | F | 31-40 | Non-functioning | 20 | c.3139C>T, p.Pro1047Ser | Missense | 0 | 0 | VUS (PM2, BP6) | - |
| *SDHA* (NM_004168.3) | 8215 | F | 19-30 | GH | 20 | c.1242C>T, p.Pro414Pro | Synonymous | 0.000024 | 0 | LB (PM2, BP6, BP7) | - |
| | 7862 | F | ≤18 | PRL | 10 | c.1409G>A, p.Ser470Asn | Missense | 0.000004 | 0 | VUS (PM2, BP4) | - |

| Gene (transcript) | Patient number (id) | Sex | Age at diagnosis (yr) | Type of adenoma (hormones produced) | Size of adenoma (mm) | Variant (nucleotide change, protein change) (a) | Effect | Allele frequency in GnomAD | Allele frequency in Portuguese controls | ACMG classification (criteria) (b) | Previous report (PMID) |
|---|---|---|---|---|---|---|---|---|---|---|---|
| | 7934 | F | 31-40 | PRL | >10 | c.1569T>C, p.Ala523Ala | Synonymous | 0.000251 | 0 | LB (PM2, BP6, BP7) | 17376234 |
| | 7685 | F | 31-40 | GH | >10 | c.1929C>T, p.Pro643Pro | Synonymous | 0.000021 | 0 | LB (PM2, BP6, BP7) | - |
| *SDHAF2* (NM_017841.2) | 8130 | F | 31-40 | ACTH | 14 | c.424G>C, p.Asp142His | Missense | 0 | 0 | VUS (PM2) | - |
| | 8092 | F | 31-40 | GH | 27 | c.496C>T, p.Arg166Cys | Missense | 0.000039 | 0 | VUS (PM2) | - |
| *SDHB* (NM_003000.3) | 7839 | M | 19-30 | Non-functioning | 17 | c.138A>G, p.Arg46Arg | Synonymous | 0.000004 | 0 | LB (PM2, BP6, BP7) | - |
| | 7887 | F | 19-30 | PRL | 20 | c.379A>C, p.Ile127Leu | Missense | 0.000004 | 0 | LP (PM1, PM2, PM5, PP2, PP3) | 23083876 |
| *SDHC* (NM_003001.5) | 8430 | M | 31-40 | PRL | 45 | c.179+10G>A | Intronic | 0.000346 | 0 | B (BS1, BS2, BP6) | - |
| *SDHD* (NM_003002.3) | 8259 | F | 19-30 | GH | >10 | c.282C>G, p.Ser94Ser | Synonymous | 0.000036 | 0 | LB (PM2, BP6, BP7) | 32688340 |
| *TP53* (NM_000546.6) | 8164 | F | 31-40 | PRL | >10 | c.31G>A, p.Glu11Lys | Missense | 0.000007 | 0 | LB (BS3, PM2, BP6) | 30352134 |
| | 7674 | F | 31-40 | GH | >10 | c.319T>C, p.Tyr107His | Missense | 0.000120 | 0 | B (BS1, BS3, BP6) | 25896519 |
| | 6907 | M | ≤18 | GH/TSH | >10 | c.467G>A, p.Arg156His | Missense | 0.000016 | 0 | VUS (BS3, PM2, PM5) | 9667734 |
| | 6907 | M | ≤18 | GH/TSH | >10 | c.582T>C, p.Leu194Leu | Synonymous | 0.000008 | 0 | LB (PM2, BP6, BP7) | 11454518 |
| | 8245 | F | ≤18 | PRL | 15 | c.642T>G, p.His214Gln | Missense | 0.000032 | 0 | VUS (PM2, PM5, BP6) | 23259501 |

| Gene (transcript) | Patient number (id) | Sex | Age at diagnosis (yr) | Type of adenoma (hormones produced) | Size of adenoma (mm) | Variant (nucleotide change, protein change) (a) | Effect | Allele frequency in GnomAD | Allele frequency in Portuguese controls | ACMG classification (criteria) (b) | Previous report (PMID) |
|---|---|---|---|---|---|---|---|---|---|---|---|
| | 8079 | F | 31-40 | ACTH | 14 | c.845G>A, p.Arg282Gln | Missense | 0.000004 | 0 | LP (BS3, PM2, PM5, PM1, PP3, PP5) | 10864200 |
| | 8360 | F | 19-30 | PRL | 15 | c.869G>A, p.Arg290His | Missense | 0.000152 | 0 | B (BS3, PM2, PM5, BP6) | 8829653 |
| *USP8* (NM_005154.5) | 7868 | F | 31-40 | GH | 18 | c.498+9A>G | Intronic | 0.000026 | 0 | VUS (PM2, BP4) | - |
| | 7889 | M | 31-40 | PRL | >40 | c.498+9A>G | Intronic | 0.000026 | 0 | VUS (PM2, BP4) | - |
| | 7656 | M | 19-30 | GH | >10 | c.1972-7A>G | Intronic | 0.000004 | 0 | VUS (PM2, BP4) | - |
| *VHL* (NM_000551.3) | 7943 | M | 19-30 | PRL | 30 | c.114C>T, p.Ser38Ser | Synonymous | 0.000039 | 0.0017 | LB (PM2, BP6, BP7) | 11536052 |
| | 6906 | F | ≤18 | PRL | >10 | c.154G>T, p.Glu52Ter | Nonsense | 0.000018 | 0 | LP (PVS1, PM2) | 21463266 |
| | 8226 | M | 19-30 | PRL | 14 | c.183C>G, p.Pro61Pro | Synonymous | 0.000762 | 0 | B (BS1, BP6, BP7) | 10408776 |
| | 7792 | M | ≤18 | TSH | 10 | c.586A>G, p.Lys196Glu | Missense | 0 | 0 | P (PM1, PM2, PM3, PP2, PP3, PP5) | 23859443 |

id, identification (anonymization code not known to anyone outside the research group); F, female; M, male; yr, years (presented as age range to protect patient privacy); ACTH, adrenocorticotropic hormone; FSH, follicle stimulating hormone; GH, growth hormone; LH, luteinizing hormone; PRL, prolactin; TSH, thyroid stimulating hormone; mm, millimeters; GnomAD, Genome Aggregation Database (v2.1.1). (a) All variants were heterozygous. (b) American College of Medical Genetics and Genomics (ACMG) classification of variants (P, pathogenic; LP, likely pathogenic; B, benign) was based on the evidence for pathogenicity [very strong (PVS1), moderate (PM1–6), or supporting (PP1–5)] or benign impact [stand-alone (BA), strong (BS1-4), or supporting (BP1-7)]. ACMG classifications were based on the web-based variant interpretation tool Franklin (Genoox Ltd, https://franklin.genoox.com/), accessed on 30 March 2024. PMID, PubMed identifier. *Publication by the authors that included the same patient.
